# A priority not guided by policy? A scooping review of policy and strategic framework on Maternal Waiting Homes

**DOI:** 10.1101/2025.01.08.25320219

**Authors:** Melvin Kunda Mwansa, Kafiswe Chimpinde, Mergan Naidoo

## Abstract

**Introduction:** Maternal Waiting Homes (MWHs) are lodgings located near health facilities where women await their delivery date and be transferred to a health facility shortly before delivery or earlier should complications arise. They are a critical strategy to improve maternal health outcomes, as they been identified as a tool for reducing maternal and perinatal mortality, especially in low-resourced countries. However, they are limited countries with deliberate policies to promote their implementation. This study aimed to review the policy and strategic framework of modern maternal waiting homes in Africa and Zambia.

**Methods:** We searched published, unpublished, and grey literature from the following sources: Google Scholar, CINAHL, PubMed, Scopus, Medline, and ResearchGate. Initial search between the 25^th^ and 27^th^ March 2024 and the final search was re-run on December 18, 2024. We used relevant synonyms and keywords (such as policy on MWHs, strategies on MWHs, National Development Plans, strategic plans, Health policies, and initiatives on MWHs in developing countries or middle- and low-income countries in the English language. The study further reviewed policies, strategies, development plans, and health strategic plans that facilitated the promotion of maternal health in Zambia.

**Results:** Database search yielded a total of ninety-four (94) items. Eighteen (18) additional items were located through hand-searching reference at the Zambia National Archives Centre. Ten (10) articles described MWH policies, strategies, and implementation guidelines from the seven (7) countries. From the Zambia National Archives Centre, nine (9) documents comprised policies, national development plans and strategic plans obtained through the national archives centre. Only one country (Timor-Leste) had a deliberate policy on maternal waiting homes. Zambia does not have a policy or strategy in place that promotes maternal waiting homes.

**Conclusion:** Like in many other African countries, In Zambia, no strategic policy has been implemented to promote maternal waiting homes. While MWHs are acknowledged in various strategic health plans and some policies, they are not consistently prioritised or adequately funded. countries, including Zambia must have MWHs policy that should employ deliberate strategies to reduce maternal and newborn morbidities and mortalities.

## Introduction

Maternity waiting homes (MWHs) are an intervention dating back to the 1950s, aimed at improving access to emergency and specialised maternity care(1). They are lodgings located near health facilities where women await their delivery date and be transferred to a health facility shortly before delivery or earlier should complications arise (2). MWHs have the potential to be used as a strategy to improve maternal health outcomes (3–6). The Zambia Demographic and Health Survey highlights (ZDHS) 2024 demonstrates that, martenal mortality ratio is 187, with the goal of achieving less than 180 maternal deaths per 100 000 live births by 2030 (7).

The Zambian government has put in intervention for improving birthing space and providing higher quality services such as emergency obstetric care post-abortion care and encouraging community-level advocacy by safe motherhood action groups. Among other interventions include the promotion of health facility deliveries by restricting traditional birth attendants from performing deliveries in communities and encouraging traditional leaders to refer their community members to health facilities for delivery (8). However, the high perinatal and maternal mortality ratio in Zambia is associated with the low accessibility to quality obstetric care as a result of delays in the decision to seek care, delays in reaching care centres, and delays in receiving care. Consequently, MWHs have been identified as a tool for reducing maternal and perinatal mortality.

The World Health Organization (WHO) recommends the establishment of MWHs close to health facilities with essential obstetric care to increase access to skilled childbirth practitioners for women living in unreachable areas or with limited access to health services (9). The purpose of MWHs is to provide a setting where pregnant women can be lodged during the final weeks of their pregnancy near a medical facility with essential obstetric services. These serve as critical interventions to improve maternal and neonatal health outcomes by providing rural pregnant women with access to skilled birth attendants and emergency services, addressing the high maternal and neonatal mortality rates that persist (2,10,11) (12) (13) (14). The MWH strategy is used to reduce maternal and perinatal mortality by improving access to skilled birth attendance and emergency obstetric care (15).

In Zambia, the Maternal Shelter Alliance (MSA) constructed twenty-two (22) maternal waiting homes in three rural provinces (Luapula, Southern and Eastern) of Zambia. Findings from the alliance report reveal that from 2016 to 2018, of all women delivering at the health facility, only 30% were utilising adjacent MWHs at intervention health facilities (Health facilities where maternal waiting homes have been constructed) (16).This indicates that around 70% of women are not utilising MWH’s, as they only opt to seek care when they recognise labour symptoms or when advised by older women or older relatives to go to the health facility to seek care despite some staying long distances from health facilities.

### Factors Influencing the Utilisation of Modern Maternal Waiting Homes

Utilisation of MWHs is influenced by factors such as the proximity of facilities, skilled care access, and maternal health benefits. Reduced distance to MWHs helps pregnant women access healthcare for the early detection of maternal complications and thereby reduce morbidity and mortality risks (17). Non-utilisation is often attributed to financial limitations, lack of family or community support, and misinformation, such as beliefs associating MWHs with witchcraft or negative cultural perceptions (18).

In Zambia, the construction of MWHs addresses significant travel barriers. Stekelenburg’s study showed that while 96% of women preferred facility-based deliveries, only 58% achieved this due to distance constraints (19). Similar findings in Ghana and Nepal underscore that long distances contribute to high rates of home deliveries, further highlighting the importance of nearby facilities for timely access ((20)-(21)). Studies across Ghana, Nigeria, and Sierra Leone also emphasise distance and cost as significant barriers to MWH usage (22).

### Barriers to Maternal Waiting Home Utilization

MWH underutilisation is often due to socioeconomic constraints. High unemployment rates in Zambia, particularly among rural women, affect MWH usage due to the costs associated with hospital-based deliveries, including transport and facility fees (23). Many women avoid MWHs due to financial constraints and the need to balance family responsibilities, such as childcare during their stay ((24) (25)). Vermeiden (26) found that women who lack financial means or who cannot leave home responsibilities are less likely to use MWHs.

Health worker attitudes also influence the utilisation of MWHs. Poor treatment by staff and lack of respect can discourage women from seeking facility-based care (27). In Malawi, a study showed that staff attitudes negatively affected women’s MWH experiences, similar to findings in Nigeria, where women left antenatal care to deliver at home due to negative staff interactions (28). However, barriers such as perceived lack of respect from staff, privacy concerns, and prohibition of traditional birthing practices may deter usage. As reported by Lori (29), women are more likely to utilise MWHs if they feel respected and well-treated.

Despite the establishment of MWHs in Zambia, there has been limited review of policies and implementation frameworks governing their promotion influence on the knowledge, attitudes and behaviours of would-be users.

Aim :

To review policies, strategies, and strategic frameworks for maternal waiting homes in African countries and Zambia in particular

### Objectives

1. Establish African countries with policies and strategies on MWHs
2. Establish Zambian policies and strategies on MWHs from 1964 to 2023
3. To establish MWH policy translations into practice

## Methods

### Data collection and tools

The Principal Investigator (PI) designed a data extraction form which was used to extract relevant information such as the topic, abstracts and objectives of the study, author, publication date, journal title, source, components addressing the topic and summary of key findings on policies on maternal waiting homes, maternal and child health in developing countries. This provided the information needed to proceed with the review. The Principal Investigator (PI) read through the reports and articles, extracting relevant information pertaining to the study and reviewed alternative successful policies over a thirty-four (34)-year period (1991 to 2024) to assess policies and strategies (reported and unreported) on maternal waiting homes.

We conducted our search by selecting published, unpublished, and grey literature and/or articles and reports from the following sources: Google Scholar, Cumulative Index to Nursing and Allied Health Literature (CINAHL), PubMed, Scopus, Medline, and Researchgate. We conducted an initial search between the 25^th^ and 27^th^ of March 2024. Where available, weekly auto alerts were requested. The final search was re-run on December 18, 2024, in the databases that did not offer auto alerts. Using relevant synonyms and keywords (such as policy on MWHs, strategies on MWHs, National Development Plans, strategic plans, Health policies, and initiatives on MWHs in developing countries or middle- and low-income countries in the English language.

We further reviewed documentation from the Zambia National Archives Centre. At the national archives centre, the review comprised of policy statements, implementation documents (including National Development Plans and National Health Strategic Plans), and published and unpublished research papers that facilitated and governed MWHs programming and construction. A total of 18 documents from the Zambia National Archives Centre were screened for inclusion, but only nine fulfilled the inclusion criteria for this study.

We conducted two training sessions with the two research assistants to equip them with systematic review skills. After an initial training session, two research assistants independently screened a 10% sample of the studies on title and abstract. Coding comparisons showed high agreement (ranging from 80% to 99% on inclusion/ exclusion criteria), therefore, the remainder of the items were independently screened. After a second training session, the two research assistants per study independently assessed the eligibility of full-text records against inclusion criteria. Discrepancies were resolved through discussion and review by the Principal Investigator (PI).

### Inclusion and exclusion criteria

Defining inclusion and exclusion criteria is imperative in any form of research or review. Specific inclusion and exclusion criteria help to clarify the intended research topic and facilitate a focused yet comprehensive search strategy. Inclusion and exclusion criteria also aid the intended audience in understanding the exact focus of the topic under study. Unpublished studies or grey literature were obtained.

| **Inclusion criteria** | **Exclusion criteria** |
| --- | --- |
| Randomised controlled trials on MWHs | Non-research-based studies, including opinion and discussion-based narratives on MWHs. |
| Non-randomised controlled trials on MWHs | Studies examining the development and implementation of MWHs, but not policy are examining the development and implementation of MWHs, but there are no policies for MWH |
| Interrupted time series, controlled before–after | Studies examing utilizutilisation and non-utilization utilisation on MWHs |
| Prospective/retrospective cohort or case–control studies on MWHs | Studies examing acceptability of MWHs |
| Studies on MWHs reported in the peer-reviewed or grey literature were included if they provided sufficient detail to assess their eligibility | Studies examining maternal mortality in developing/low-income countries with no mention of MWHs |
| Policies or policy discussions on MWHs | Studies and intervention with no policy guidance and/or direction |
| Strategies on MWHs | Studies not available in full-text |
| Implementation guidelines on MWHs | Studies not freely accessible on the World Wide Web or, through electronic databases |
| Zambian policies on maternal health strategies from 1965 to 2023 |  |
| Zambian national development plans/documents from 1965 to 2023 |  |
| The language was restricted to English |  |

### Data analysis techniques

The policy development plans and strategic framework on MWHs in Africa, and particularly in Zambia, were analysed using a qualitative method called framework analysis. Data was cleaned thoroughly through organising and keeping track of all raw records. Raw records included raw data files (policies, national development, and strategic plans), as well as both published and unpublished materials from the national archives centre and government departments. Summarised data provided a structure in which the researcher systematically reduced the data to tell a story and analysed it by case and code as guided by the framework method. The data cleaning process undertook levels of reviews, with the research assistant conducting the first review by gathering all documents and counterchecking the themes. At the same time, the PI also reviewed all documents and compared it with the synthesised material in the extraction form, ensuring that the content was correct.

The mixed method appraisal tool version 18 was used. The MMAT is designed for the appraisal stage of systematic mixed studies reviews, i.e., reviews that include qualitative, quantitative and mixed methods studies. It permits to appraise the methodological quality of five categories to studies: qualitative research, randomized controlled trials, non-randomized studies, quantitative descriptive studies, and mixed methods studies (30). The MMAT is used to appraise the quality of empirical studies, i.e., primary research based on experiment, observation or simulation

### Ethical Considerations

The study was approved by the University of KwaZulu Natal - Health and Social Sciences Research Ethics Centre (HSSREC) reference number HSSREC/00002978/2021 and the National Health Research Authority (NHRA) reference number NHRA00014/17/08/2021.

## Results

African countries with policies and strategies on MWHs database search yielded a total of ninety-four (94) items (Figure 1). Eighteen (18) additional items were located through hand-searching references at the Zambia National Archives Centre. Articles were included in this review if they met the set inclusion criteria. Searching ended when there were no new references being found in the reference lists, and the citation of references became circular. In total, forty-eight (48)articles and/or records were identified for full-text evaluation. Of these, thirteen (13) articles and/or records did not meet the inclusion criteria. The articles and records were further assessed against all elements of the inclusion criteria. Thirteen (13) articles and/or documents were included, as highlighted in Table 1 below. Study characteristics included policy and strategic documents and articles published between 1991 and 2024 from the following countries: Ethiopia (n =1), Eriteria (n=1), Tanzania (n= 1),) Timor-Leste (n=1), Uganda (n=1), Ghana (n=2), Lesotho (n=1) and Zambia (n-9) (Figure 2). Ten (10) reports described MWH policies, strategies, and implementation guidelines from the seven (7) countries highlighted above

**Figure 1.**
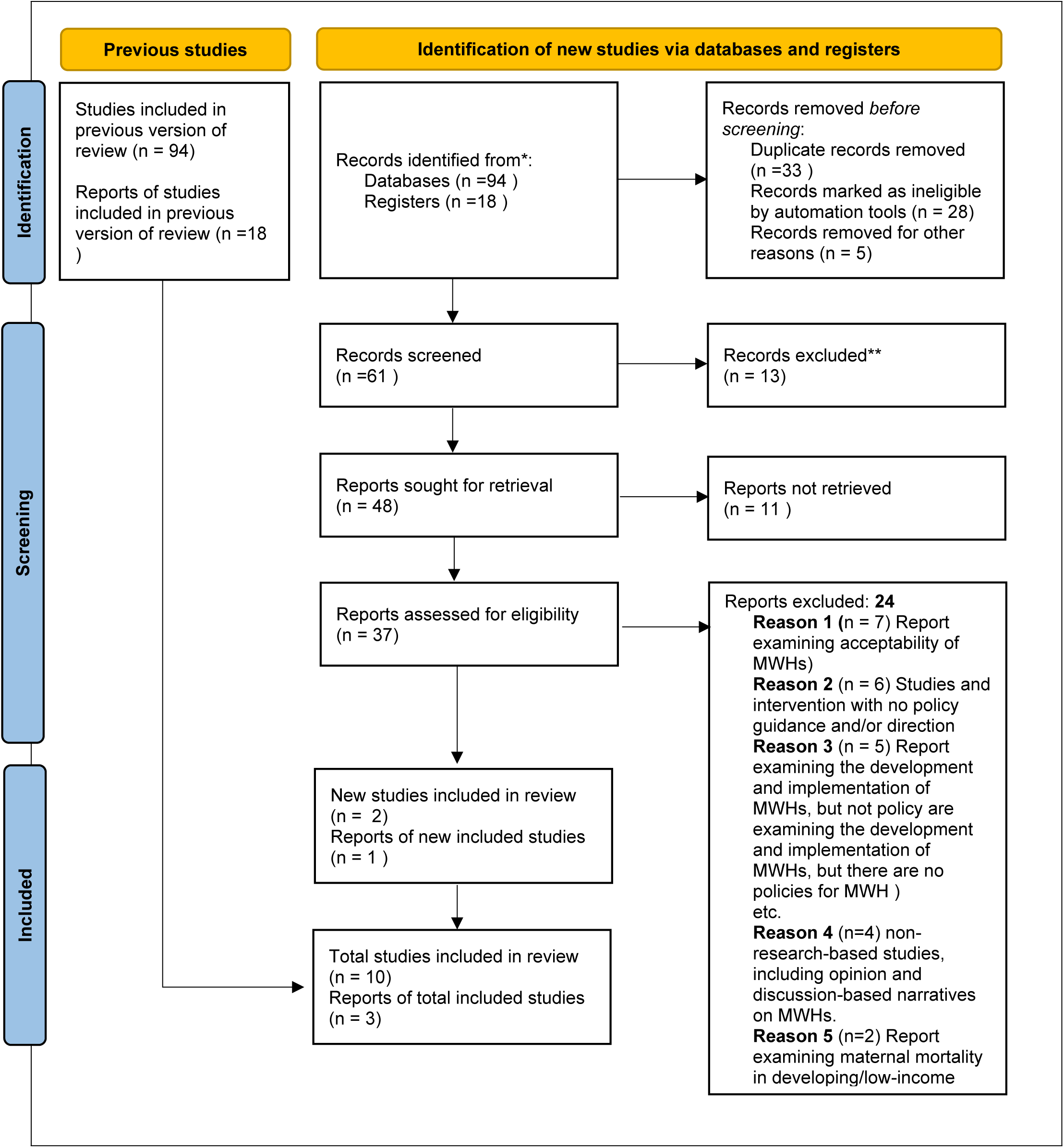
updated PRISMA 2020 flow diagram for searches of databases and registers (studies and documentation)

**Table 1:**
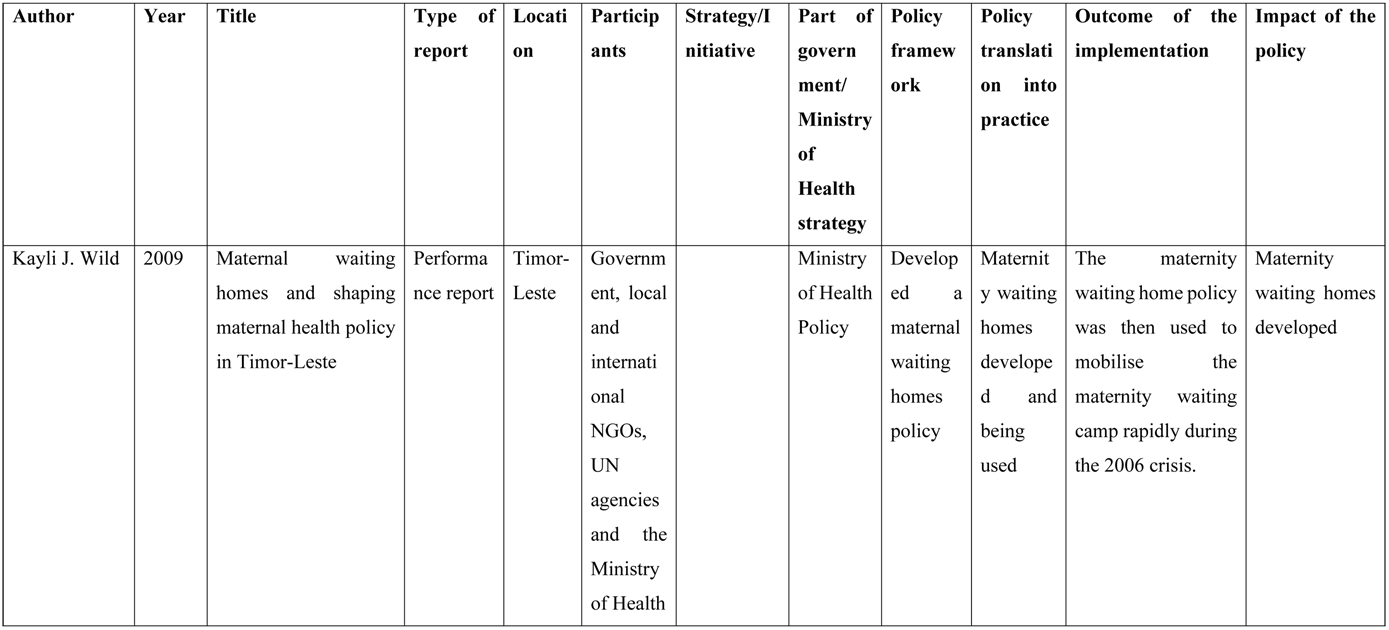
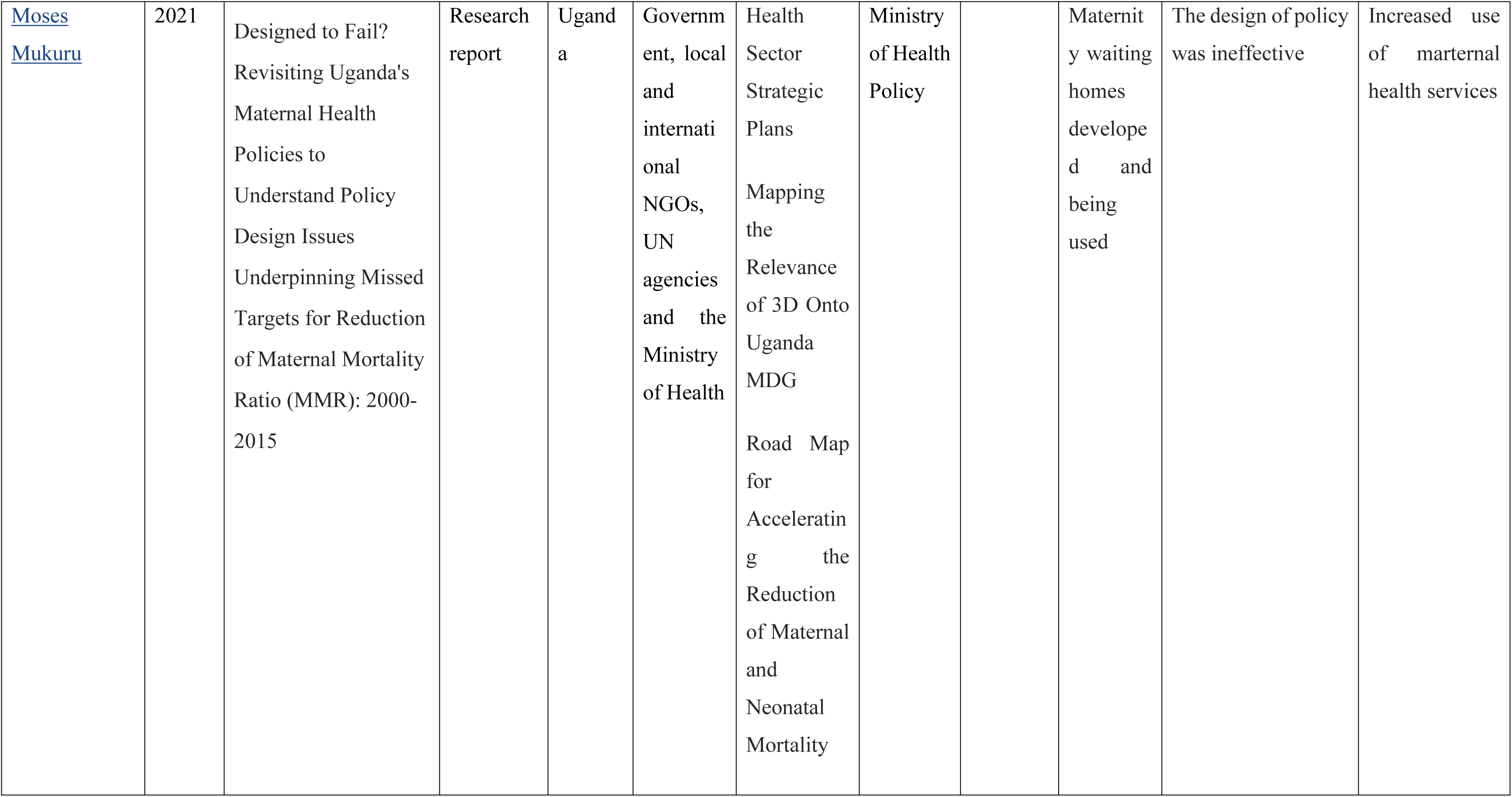
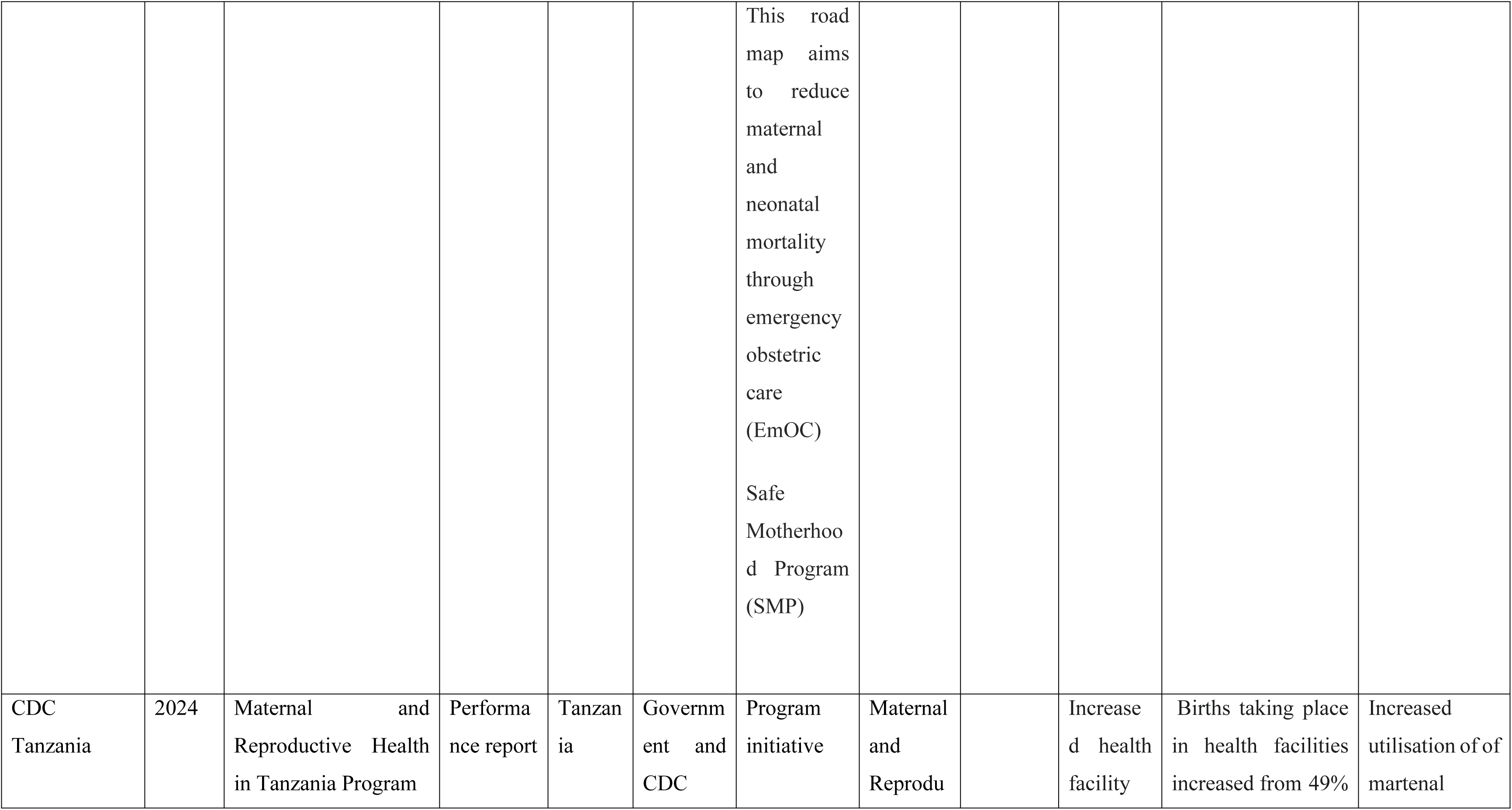
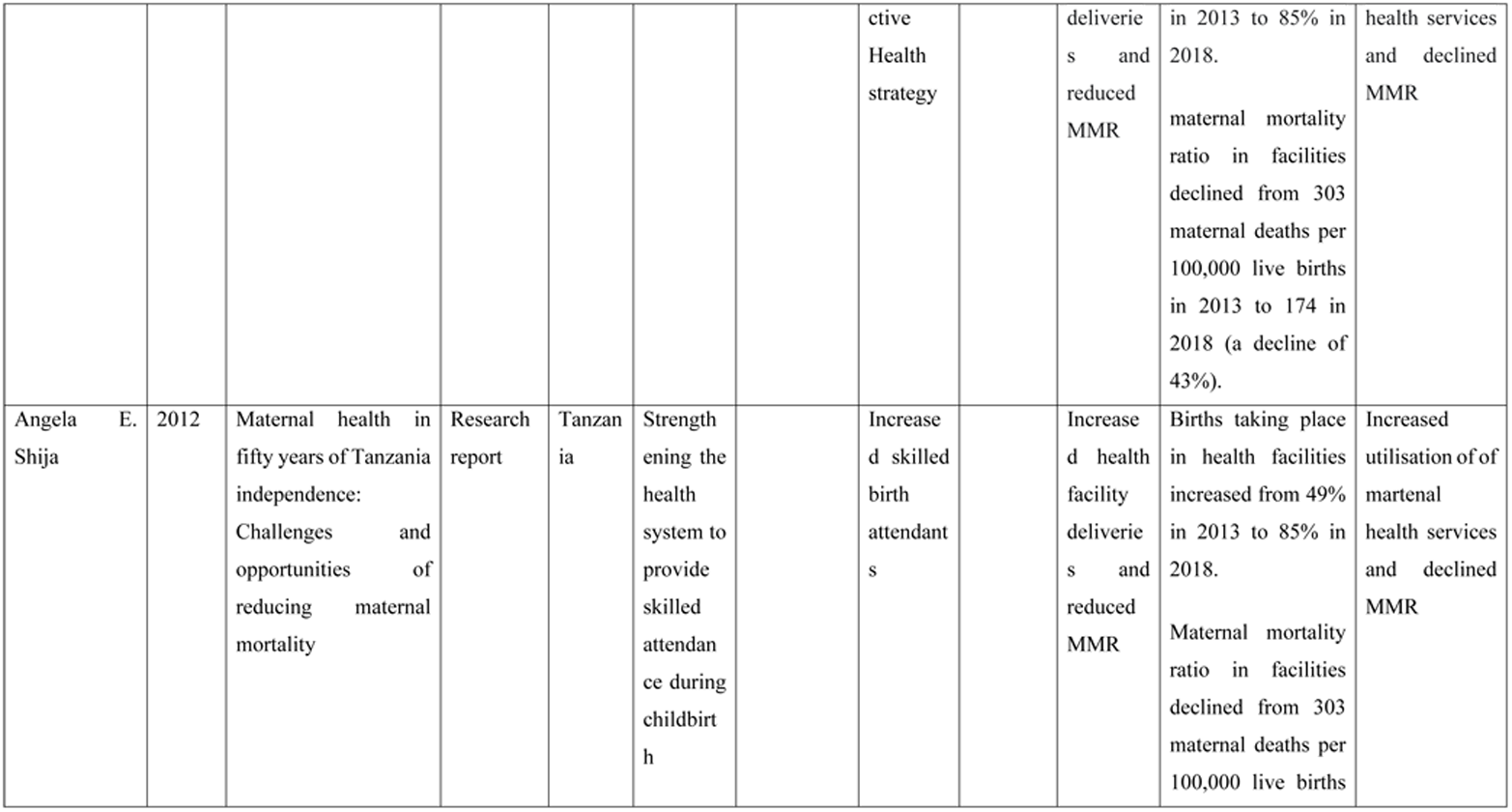
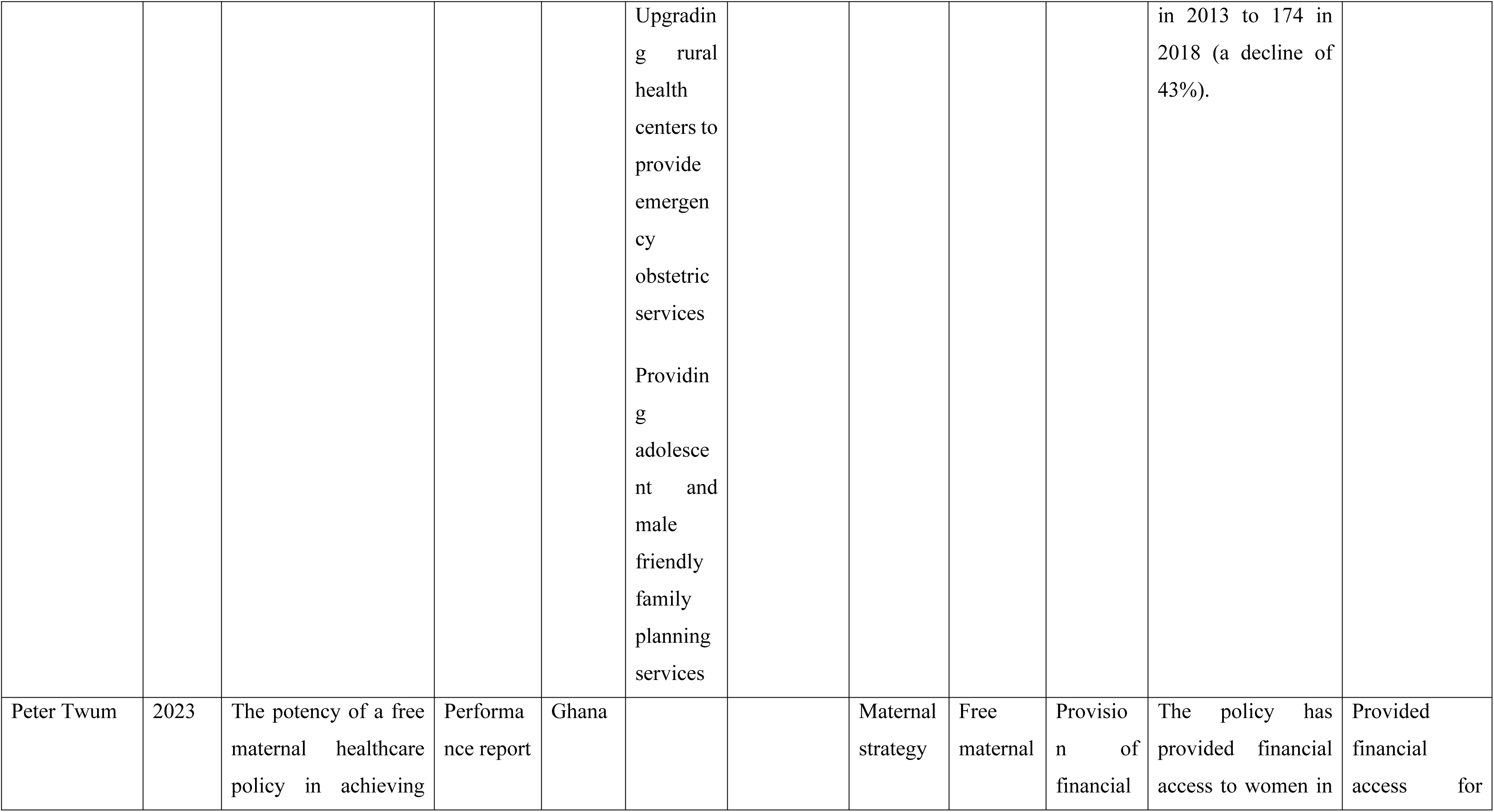
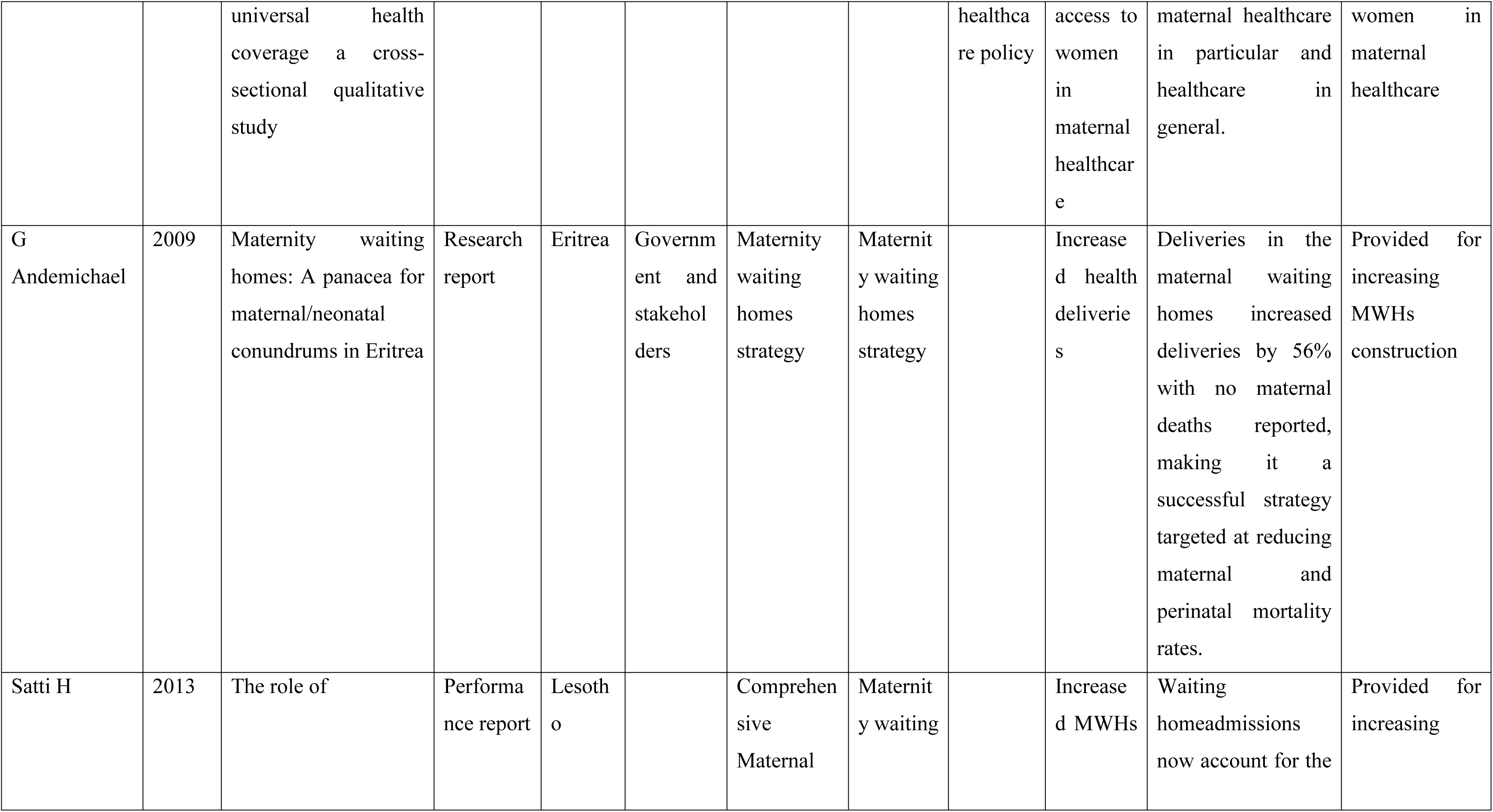
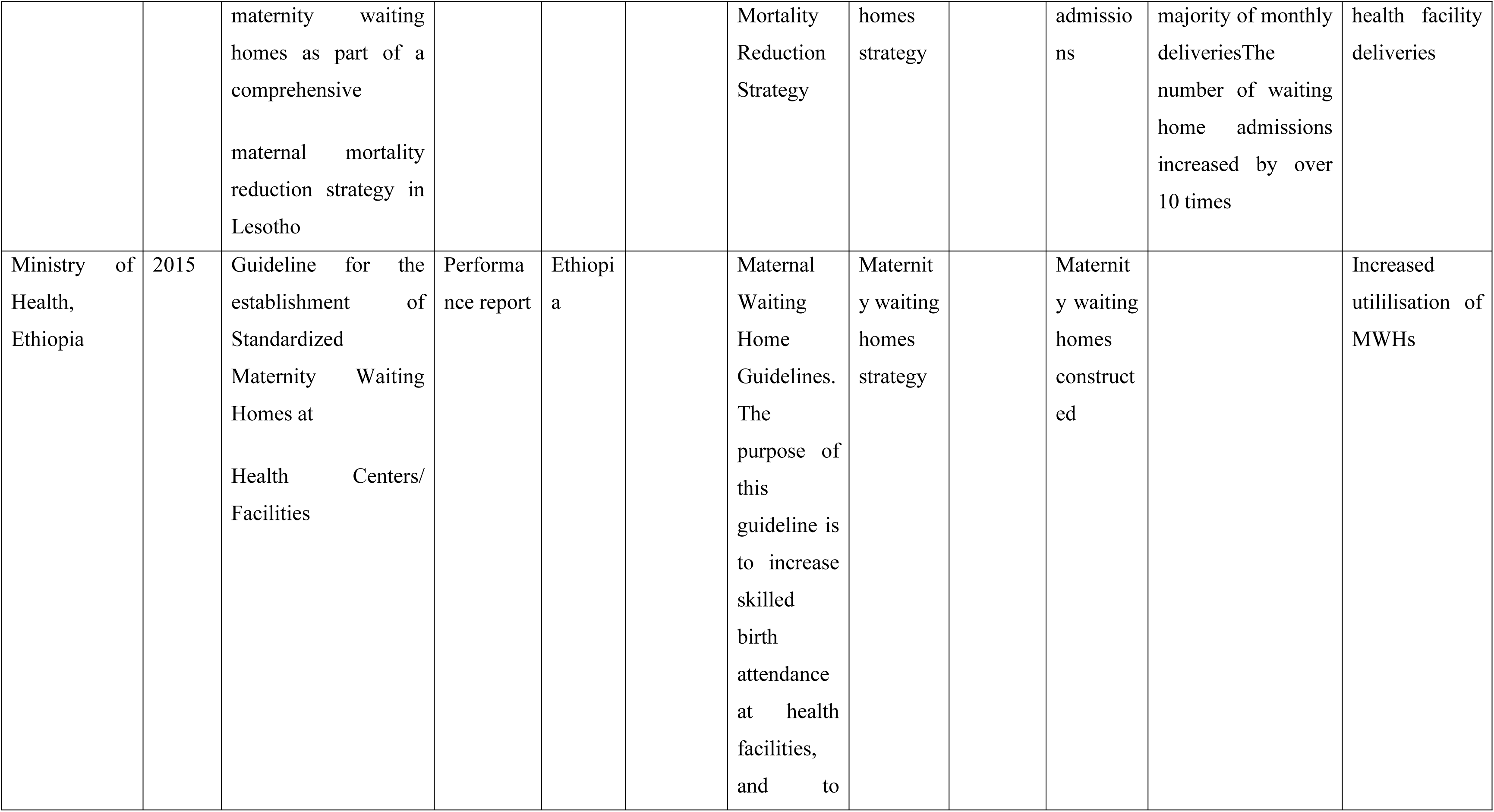
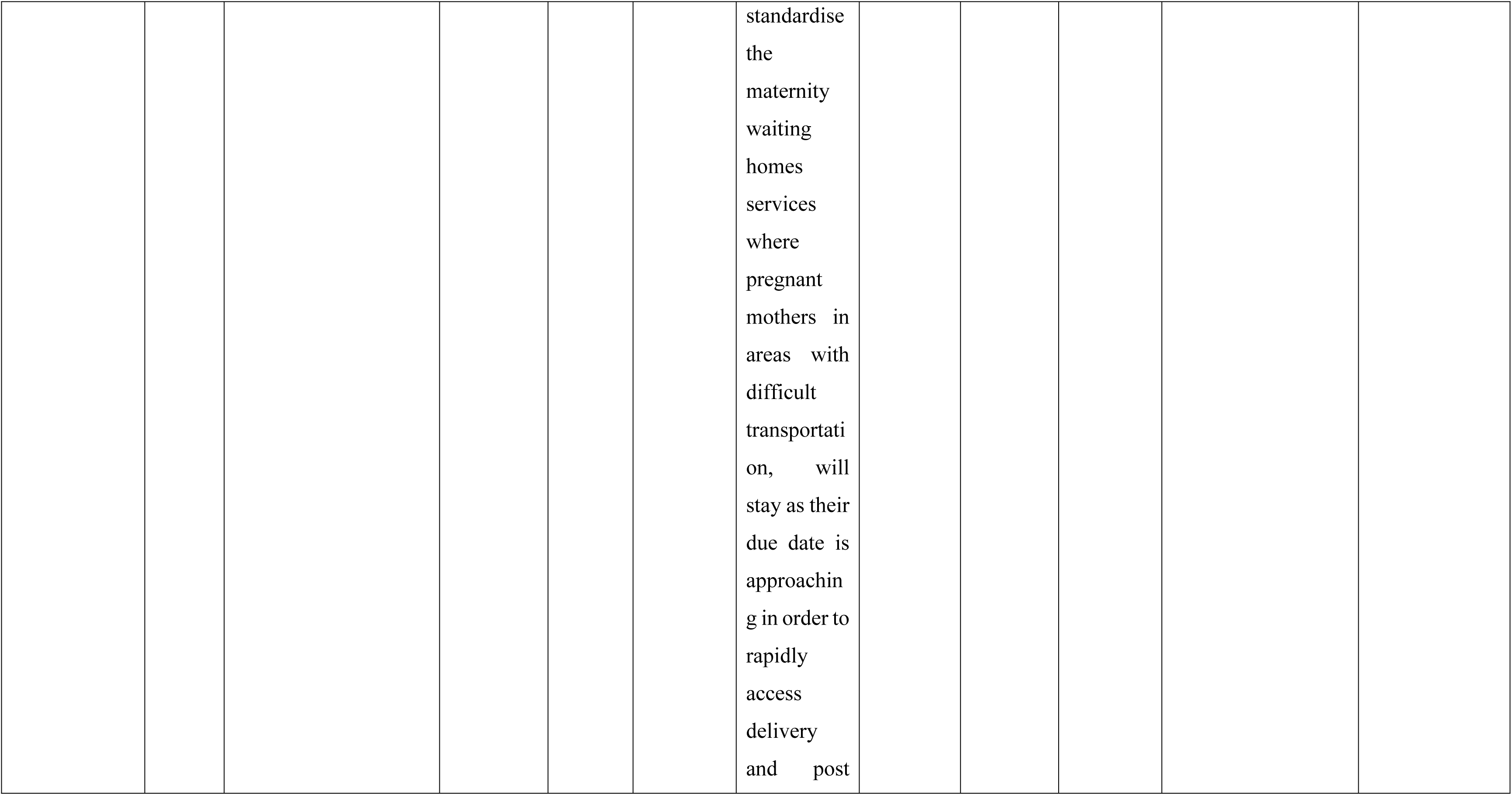
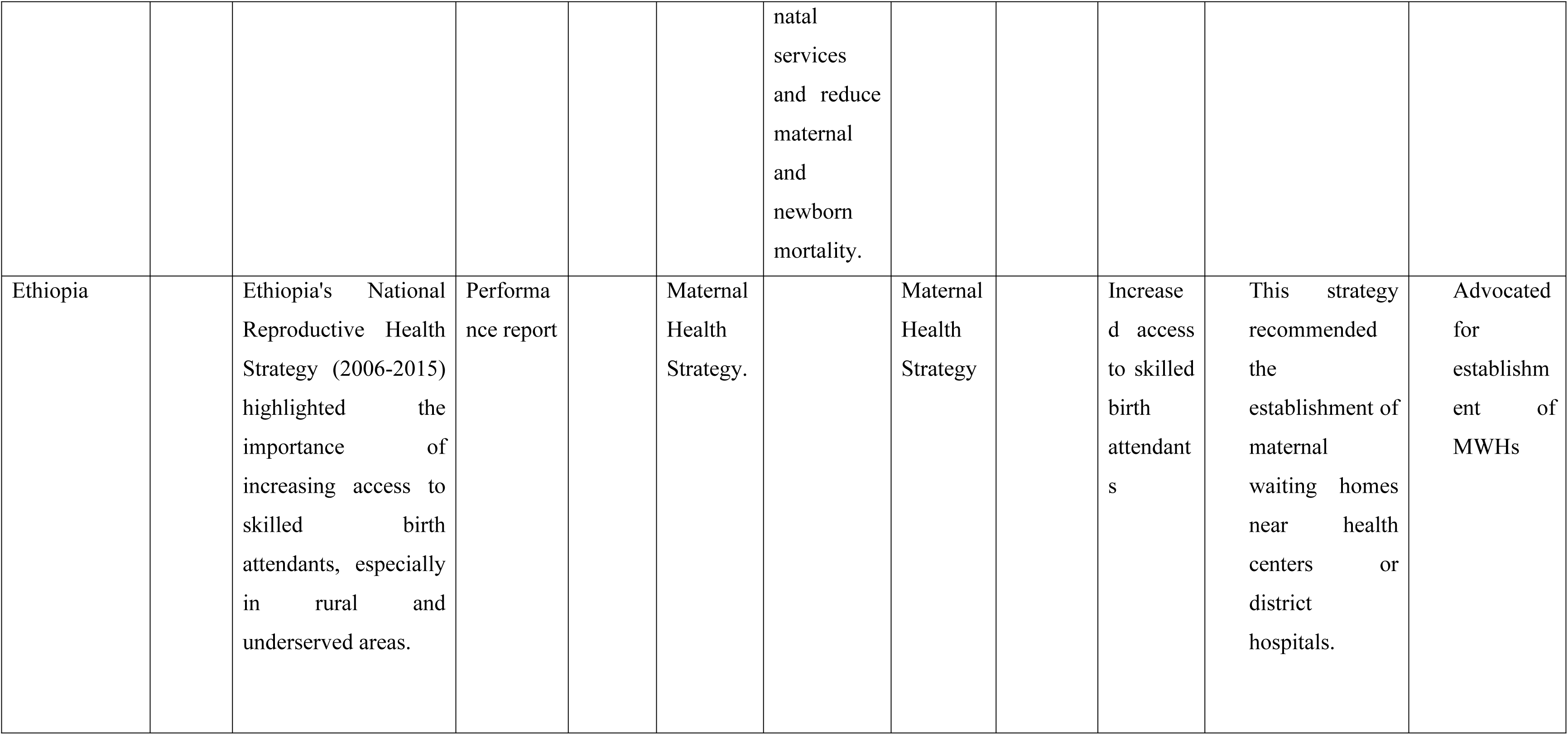
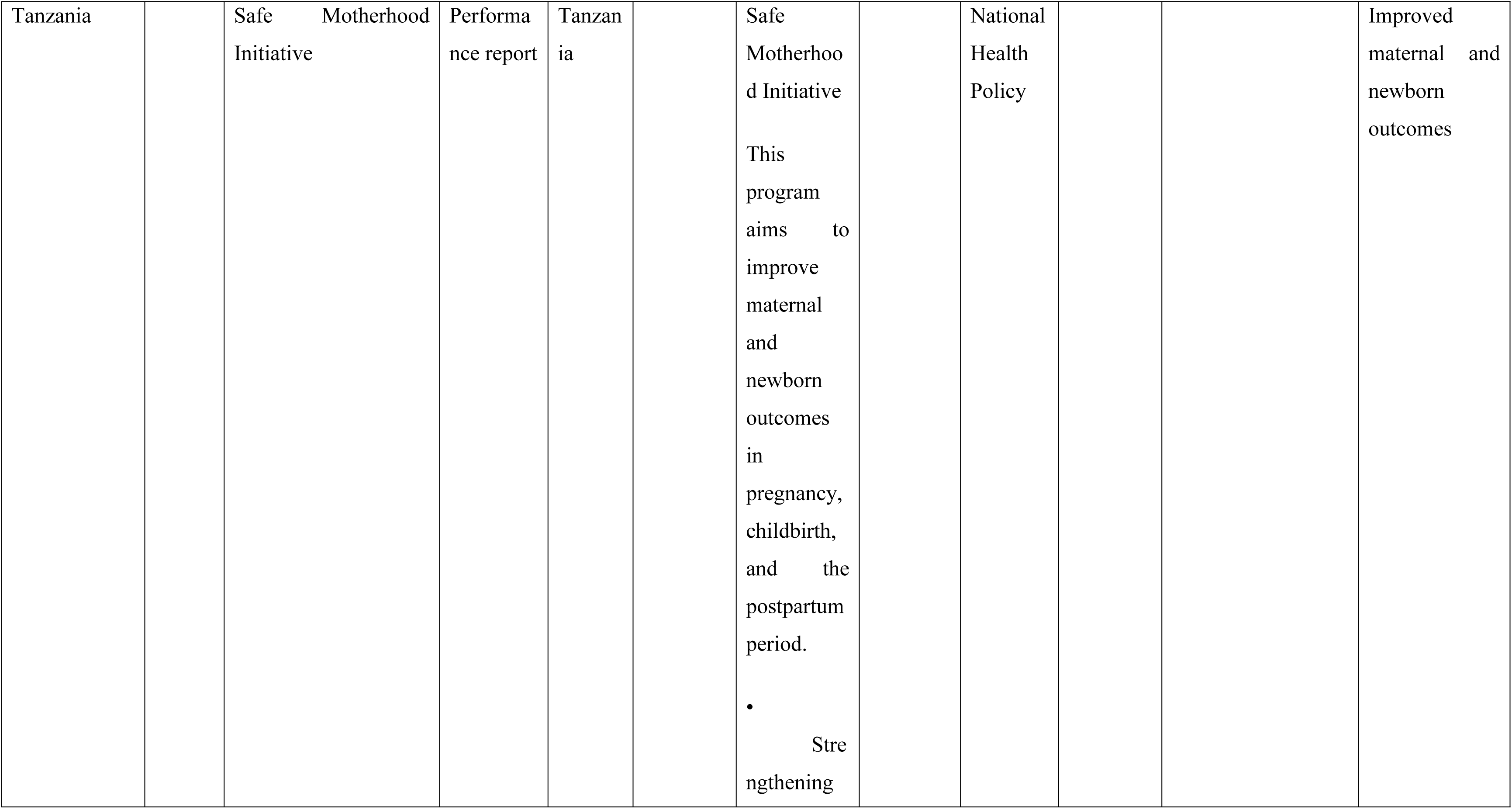
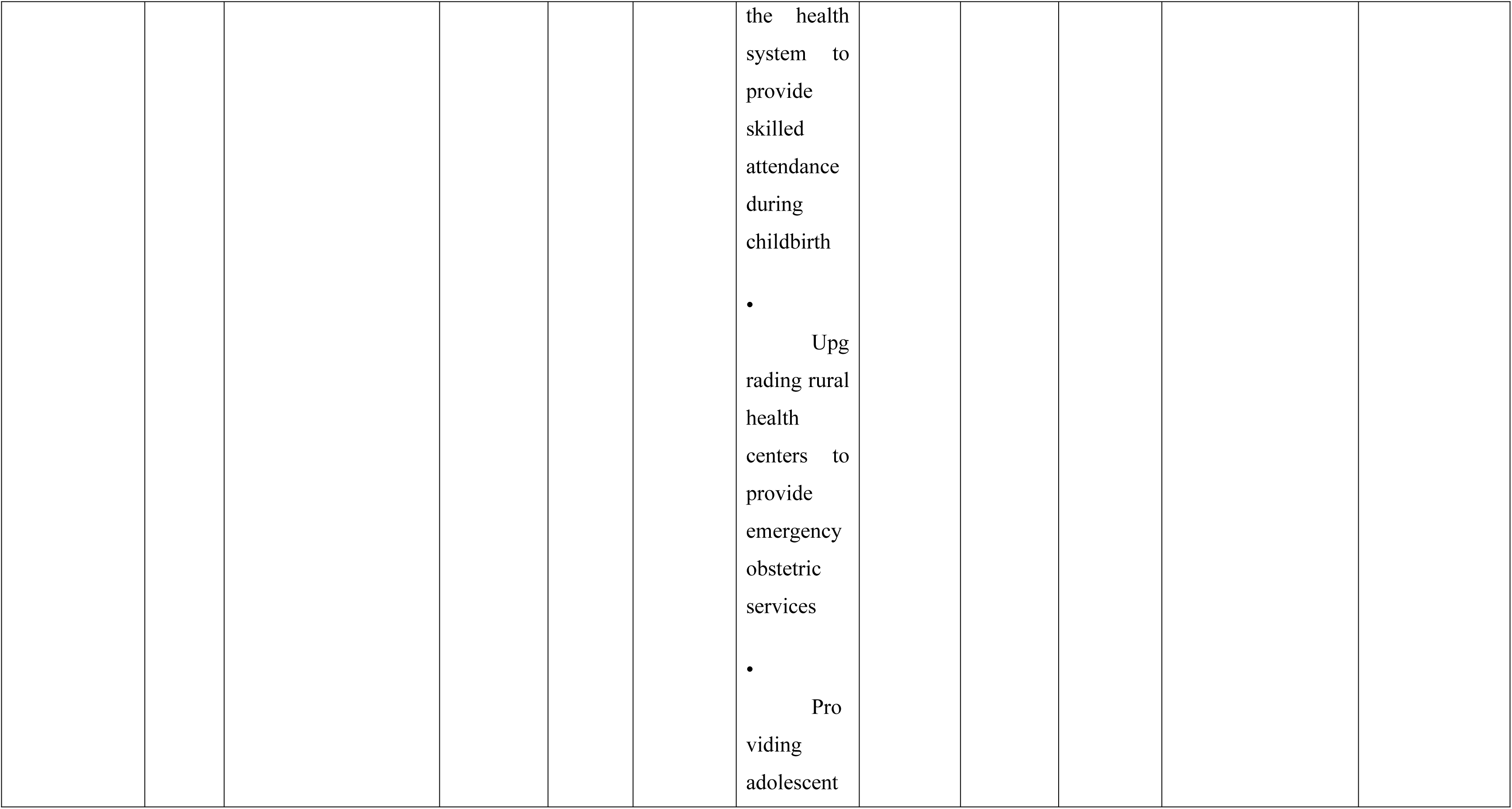
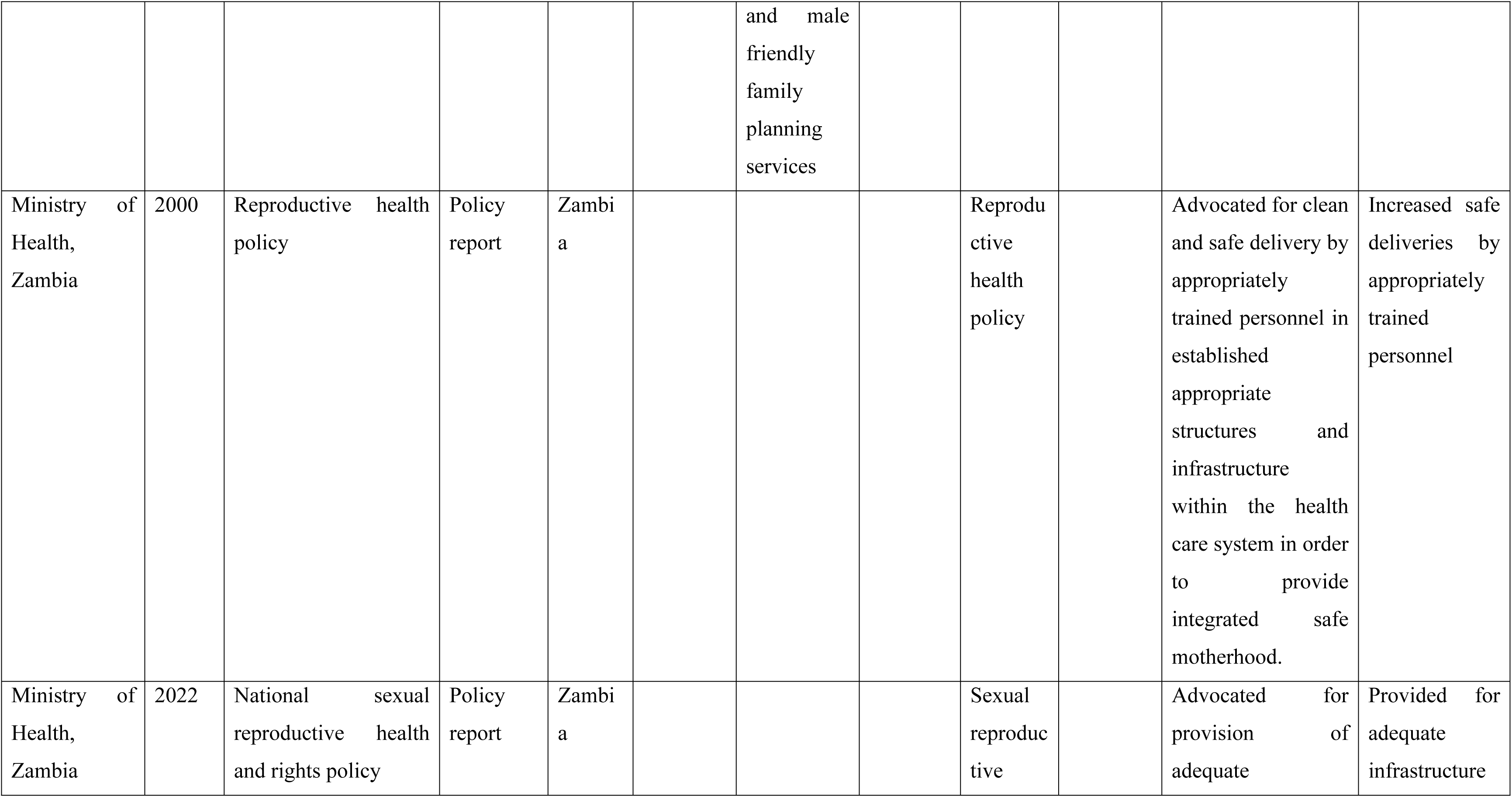
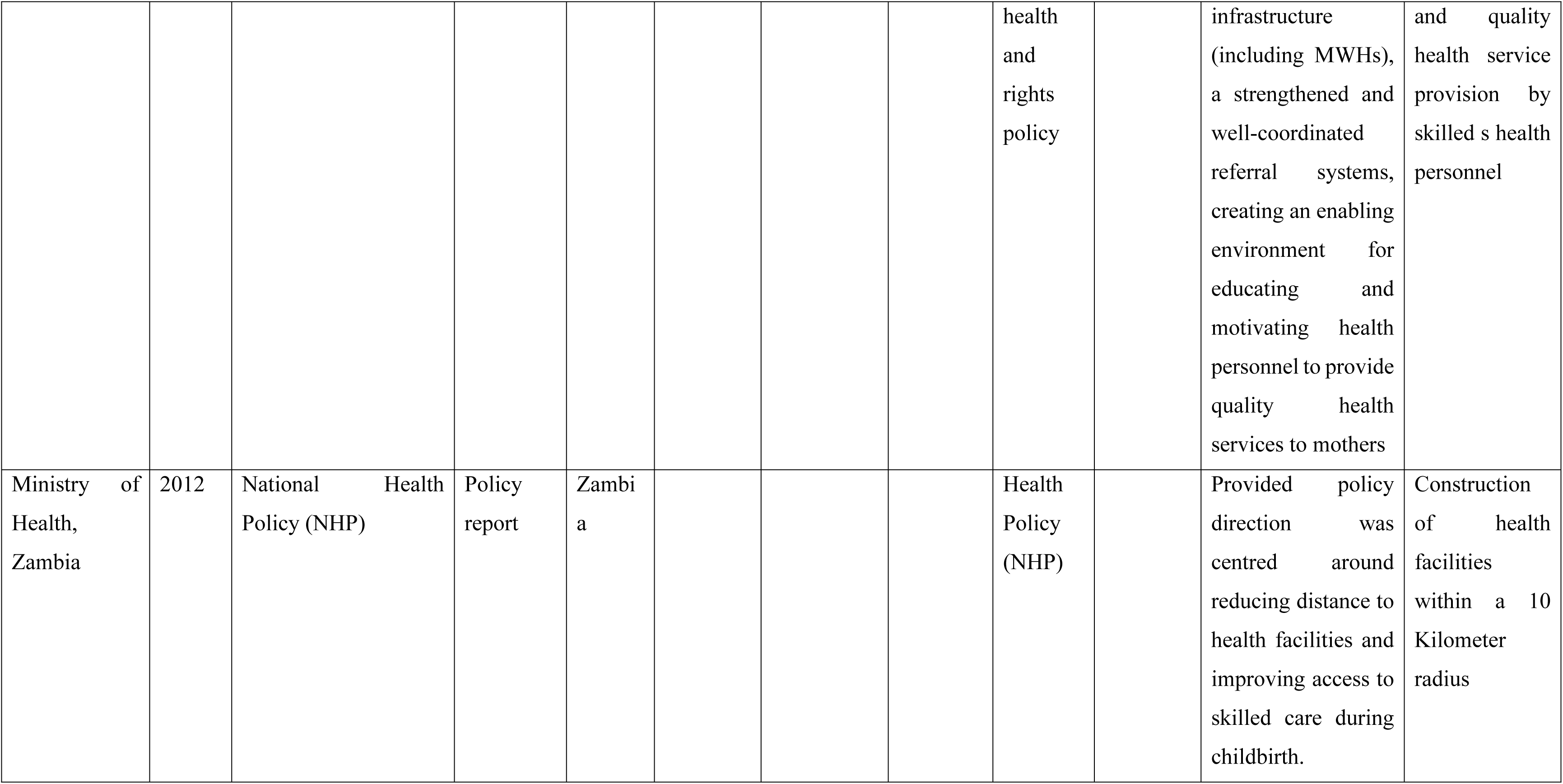
Selected studies and reports on policy and strategic frameworks on modern maternal waiting homes.

The review review shows that in Africa,, only Timor-Leste had a specific MWH policy (31), whilst the seven other countries had employed strategies and/or guidelines on MWHs. The Ugandan government introduced fourteen separate policy instruments over 15 years with the goal of reducing maternal mortality. Tanzanian government introduced three (3) strategies aimed at improving maternal health. The specific strategies included; strengthening the health system to provide skilled attendance during childbirth, upgrading rural health centres to provide emergency obstetric services and providing adolescent and male friendly family planning services (38).

In Ghana, the government introduced free maternal healthcare policy (FMHCP)) under the national health insurance scheme (NHIS) in 2008 while the Eritrean governement implemented a maternity waiting homes strategy in 2007. The government in Lesotho introduced comprehensive maternal mortality reduction strategy. Similarly, in 2015, the Ethopian government introduced guidelines for the establishment of standardised maternity waiting homes at health centres/ facilities.

### MWH policy translations into practice

In Timor-Leste, the maternity waiting home policy was used to rapidly mobilise the maternity waiting camps during the 2006 crisis (32). In Tanzania, the introduced initiatives and strategies resulted in increased health facility deliveries (49% in 2013 to 85% in 2018) and a 43% declining maternal mortality ratio in health facilities (303 maternal deaths per 100,000 live births in 2013 to 174 in 2018) (33). The Ghanian policy promoted and increased health facility utilisation and deliveries being attended to by skilled personnel (34).

The study established that the Eritrean strategy resulted in increased deliveries in the maternal waiting homes by 56% with no maternal deaths reported, making it a successful strategy targeted at reducing maternal and perinatal mortality rates (5). In Lesotho, MWH admissions accounted for the majority of monthly deliveries and the number of waiting home admissions increased by over 10 times (35). The Ethopian Government’s strategy recommended the establishment of maternal waiting homes near health canters or district hospitals (36).

### Zambian policies and strategies on MWHs from 1964 to 2023

Three (3) national policies highlighted promotion of maternal outcomes through construction of MWHs, with two national development plans, two national health strategic plans, one initiative and one act indirectly promoting maternal health outcomes and MWHs, especially in rural areas;

1. The Reproductive Health Policy of 2000 aimed at ensuring the provision of quality, integrated safe motherhood services were accessible, acceptable, and free at all levels of the health care system. The policy advocated for clean and safe delivery by appropriately trained personnel in established appropriate structures and infrastructure within the health care system to provide integrated safe motherhood. The policy further encouraged postnatal care attendance for early detection of risks of maternal death after delivery (37).
2. The National Health Policy (NHP) of 2012 resonated with improving maternal and child health outcome with the goal of goal improving maternal health and reduce maternal and neonatal mortality. In the NHP, policy direction was centred around reducing distance to health facilities and improving access to skilled care during childbirth. The policy viewed MWHs as part of broader rural healthcare delivery improvements and strongly acknowledge their potential to improve access to timely care (38).
3. National sexual reproductive health and rights policy of 2022 was envisioned at making informed reproductive health choices by 2030 with a goal of reducing mortality and morbidity among women during pregnancy, childbirth, and postnatal period through implementation of combination of health and health related measures. The policy advocated for provision of adequate infrastructure (including MWHs), a strengthened and well-coordinated referral systems, creating an enabling environment for educating and motivating health personnel to provide quality health services to mothers, women, young women, and families.

The study established other national development plans, strategic plans, strategies and acts developed with the aim of improving livelihoods of Zambians and improve maternal health. Zambia has implemented eight national development plans from independence in 1964 to date. The review established that only two national development plans resonate with promotion of maternal and child health interventions. The Fifth National Development Plan implemented between 2006-2010 and the seventh national development plan (7NDP). The fifth national development plan 2006-2010, was in line with vision 2030, as it aimed at having equitable access to quality health care for all by 2030 and expanding access to Maternal, New-born and Child Health services including immunisation, Family Planning, safe delivery, and Basic Emergency Obstetric Care with special focus on under-served areas and the vulnerable population (39). Despite the 7NDP the development plan aiming at improved maternal and child health, the focus was not placed on MWHs as an intervention but increased attention to increasing proportion of skilled attended deliveries (40).

Zambia has developed and implemented six National Health Strategic Plans (NHSP) that cover a period of five years. The review indicates that, of the six NHSP developed and implemented only two (2017 - 2021 NHSP) and (2022 – 2026 NHSP) were aligned with improvements in maternal and child health. The (2017 - 2021 NHSP) was centred on improving maternal and child health with the goal of improving the health status of people in Zambia in order to contribute to increased productivity and socioeconomic development with a vision of a Nation of Healthy and productive people (41). The plan aimed at reducing MMR from 398 deaths per 100,000 to 162 deaths per 100, 000 live births and reducing under-five mortality rate from 75 to 56 deaths per 1,000 live births (41).

National Health Strategic plan 2022-2026 was aimed at reducing maternal mortality raion (MMR) from 278 deaths to less than 100 deaths per 100, 000 live births and neonatal Mortality Rate from 27 deaths per 1000 live births to 12 deaths per 1000 live births (42).

The roadmap for Accelerating Reduction of Maternal, New-born and Child Mortality, 2013-2016 was aimed at attaining the set 2016 targets through accelerated reduction of maternal, new-born and childhood morbidity and mortality. The roadmap was focused on reducing maternal mortality from 591 (in 2007) to 159 per 100,000 live births by 2016, and further reducing neonatal mortality from 34 to 20 per 1,000 live births by 2016 (43). The Gender equity and equality act of 2015 promoted gender equity in health access. The act supported implicitly the integration of MWHs in maternal health strategies (44).

Despite all highlighted government development plans, strategic documents and policies focusing on maternal and child health improvements. No strategic policy has been implemented to promote maternal waiting homes in Zambia. Instead, general maternal health interventions such as safe motherhood services, family planning, and skilled delivery attendance were prioritised.

## Discussion

From the review, only Timor-Leste had developed a specific maternal waiting home (MWH) policy, whereas seven other countries relied on strategies or guidelines. In Timor-Leste, the government introduced a MWH policy in 2005 and collaborated with stakeholders, including community beneficiaries, to implement these homes (33). Networks established between expert advisors and junior bureaucrats influenced the development of the policy content, which primarily aligned with WHO guidelines (34). Working groups and national meetings clarified policy content and built consensus (35). District health and village leaders played a pivotal role in bridging the national agenda with peripheral services and communities. The MWH policy proved instrumental during the 2006 crisis, enabling the rapid establishment of maternity waiting camps (36).

In Uganda, the government introduced 14 separate policy instruments over 15 years to reduce maternal mortality. By the end of the Millennium Development Goals (MDG) period in 2015, 87.5% of interventions addressing the “three delays” were covered. However, a lack of coherence and consistency among instruments was evident (37). Mururu argues that ineffective policy design, despite strategic plans and mapping of the “three delays” onto the Uganda MDG agenda, hindered the reduction of maternal mortality (37).

The Tanzanian government, with support from the Centers for Disease Control (CDC) and other stakeholders, implemented a maternal and reproductive health initiative to improve maternal health. Shija highlights three specific strategies: strengthening health systems to provide skilled childbirth attendance, upgrading rural health centers to offer emergency obstetric services, and providing adolescent- and male-friendly family planning services (38). These efforts led to increased health facility deliveries (49% in 2013 to 85% in 2018) and a 43% decline in the maternal mortality ratio in facilities (303 per 100,000 live births in 2013 to 174 in 2018) (39). Strengthening community participation and empowering women were recommended as essential strategies for further reducing maternal deaths (38).

In Ghana, the government introduced the Free Maternal Healthcare Policy (FMHCP) under the National Health Insurance Scheme (NHIS) in 2008, aiming for universal health coverage (40). The policy improved access to maternal healthcare, increased facility-based deliveries attended by skilled personnel and contributed to better maternal and neonatal health outcomes (41).

Eritrea implemented an MWH strategy in 2007, which resulted in a 56% increase in deliveries at these facilities. No maternal deaths were reported, highlighting its success in reducing maternal and perinatal mortality (5).

Lesotho introduced a comprehensive maternal mortality reduction strategy that included MWHs. The strategy led to a significant increase in waiting home admissions, accounting for the majority of monthly deliveries. Facility-based deliveries rose from an average of 3.8 per month before implementation to 18.0 within two years, with coverage increasing from 14% to 64% (42, 43). The government continues to develop strategies to reduce maternal mortality by increasing skilled practitioner coverage.

In Ethiopia, the government introduced the National Reproductive Health Strategy (2006–2015), which emphasised increasing access to skilled birth attendants in rural and underserved areas. In 2015, standardised guidelines for MWH establishment were introduced to improve skilled birth attendance and reduce maternal and neonatal mortality (44, 45).

In Zambia, policies focused on broader maternal health goals rather than explicitly prioritising MWHs. While frameworks like the National Health Strategic Plan (NHSP) and the Reproductive, Maternal, Newborn, Child, and Adolescent Health and Nutrition (RMNCAH-N) strategy mention expanding MWHs, significant gaps in financial and regulatory frameworks hinder their sustainability. Poor infrastructure and referral systems in rural areas further limit their effectiveness. Developing specific policies and resource allocation mechanisms is essential for MWHs to realise their potential.

### Recommendations

- Develop specific financial and implementation frameworks to improve the sustainability of MWHs.
- Integrate MWHs into maternal health strategies through outreach services and community education.
- Increase investment in rural education programs and transportation support for expectant mothers.
- Allocate more resources toward MWH infrastructure and staffing.
- Strengthen community outreach on the importance of skilled delivery services and provide subsidies for MWH-related costs.

### Limitations

The study faced logistical challenges in accessing archival documents from other countries. The review was limited to unpublished data from Zambia.

## Conclusion

In Zambia, as in many other African countries, maternity waiting homes (MWHs) remain an underutilised but critical strategy for improving maternal and neonatal health outcomes. Despite their inclusion in various health plans and strategies, a lack of consistent prioritisation, robust policies, and adequate funding limits their full potential. The experiences of countries such as Eritrea, Ethiopia, and Lesotho underscore the transformative impact of targeted MWH strategies when coupled with political commitment, community involvement, and adequate resource allocation.

Eritrea’s success in significantly increasing facility-based deliveries with no maternal deaths demonstrates how well-planned and executed MWH initiatives can lead to substantial improvements in maternal and perinatal health. Similarly, Lesotho’s comprehensive maternal mortality reduction strategy, which included MWHs, resulted in dramatic increases in facility-based deliveries and coverage. These examples highlight the importance of integrating MWHs into broader maternal health strategies and ensuring their alignment with national health goals.

Tanzania and Ethiopia provide further insights into the role of strategic frameworks and stakeholder collaboration. Tanzania’s multifaceted approach—strengthening health systems, upgrading facilities, and expanding access to family planning—led to a notable decline in maternal mortality. Ethiopia’s establishment of standardised guidelines for MWHs showcases the critical role of government leadership and structured policy support in improving maternal health services.

The Ugandan experience, however, illustrates the risks of fragmented policies and inconsistent implementation. While Uganda made significant strides in addressing the “three delays,” the lack of coherence among policy instruments undermined efforts to reduce maternal mortality.

These diverse experiences offer valuable lessons for Zambia and other countries seeking to leverage MWHs to improve maternal health outcomes. For MWHs to play a more significant role, governments must implement specific policy directives backed by high-level political will, sustainable funding, and community engagement. Strengthened regulatory frameworks, increased investment in infrastructure, and coordinated efforts across government sectors and stakeholders are essential. Furthermore, aligning MWH strategies with broader initiatives such as universal health coverage and community empowerment can maximise their impact.

Ultimately, the successful implementation of MWHs across Africa requires a holistic approach that addresses the social determinants of health, improves transportation and referral systems, and ensures access to skilled care for all pregnant women. By learning from successful models in Eritrea, Lesotho, and Ethiopia, and addressing gaps identified in countries like Uganda and Zambia, African nations can build stronger health systems capable of reducing maternal and neonatal mortality and achieving Sustainable Development Goals.

## Data Availability

Data is avaible

